# A systematic review of quantitative EEG findings in Long COVID, Fibromyalgia and Chronic Fatigue Syndrome

**DOI:** 10.1101/2023.11.06.23298171

**Authors:** Bárbara Silva-Passadouro, Arnas Tamasauskas, Omar Khoja, Alexander J. Casson, Ioannis Delis, Christopher Brown, Manoj Sivan

**Affiliations:** Leeds Institute of Rheumatology and Musculoskeletal Medicine, School of Medicine, University of Leeds, Leeds, UK; Institute of Life Course and Medical Sciences, Faculty of Health and Life Sciences, University of Liverpool, Liverpool, UK; Department of Electrical and Electronic Engineering, University of Manchester, Manchester, UK; School of Biomedical Sciences, Faculty of Biological Sciences, University of Leeds, Leeds, UK; Department of Psychology, Institute of Population Health, University of Liverpool, Liverpool, UK; National Demonstration Centre in Rehabilitation Medicine, Leeds Teaching Hospitals NHS Trust, Leeds, UK

**Keywords:** Electroencephalography, Central sensitisation, COVID-19, Post-Covid Syndrome (PCS), Post Acute Covid Syndrome (PACS), Post-Covid Condition (PCC)

## Abstract

Long COVID (LC) is a multisymptom clinical syndrome with similarities to Fibromyalgia Syndrome (FMS) and Chronic Fatigue Syndrome/Myalgic Encephalomyelitis (CFS/ME). All these conditions are believed to be associated with centrally driven mechanisms such as central sensitisation. There is a lack of consensus on quantitative EEG (qEEG) changes observed in these conditions. This review aims to synthesise and appraise the literature on resting-state qEEG in LC, FMS and CFS/ME, to help uncover possible mechanisms of central sensitisation in these similar clinical syndromes. A systematic search of MEDLINE, Embase, CINHAL, PsycINFO and Web of Science databases for articles published between December 1994 and September 2023 was performed. Following screening for predetermined selection criteria and out of the initial 2510 studies identified, 17 articles were retrieved that met all the inclusion criteria, particularly of assessing qEEG changes in one of the three conditions compared to healthy controls. All studies scored moderate to high quality on the Newcastle-Ottawa scale. There was a general trend for decreased low-frequency EEG band activity (delta, theta, and alpha) and increased high-frequency EEG beta activity in FMS, whereas an opposite trend was found in CFS/ME. The limited LC studies included in this review focused mainly on cognitive impairments and showed mixed findings not consistent with patterns seen in FMS and CFS/ME. Further research is required to explore whether there are phenotypes within LC that have EEG signatures similar to FMS or CFS/ME. This could inform identification of reliable diagnostic markers and possible targets for neuromodulation therapies.

## 1. Introduction

Long COVID (LC) is a new clinical syndrome posing a novel challenge to healthcare services globally. According to the National Institute for Health and Care Excellence (NICE)’s guidelines, LC refers to persistent symptoms lasting more than four weeks following acute coronavirus disease 2019 (COVID-19) infection (NICE, 2022). NICE also defines Post-COVID Syndrome (PCS) as a subtype of LC when symptoms persist more than 12 weeks. PCS is also called Post-COVID Condition (PCC) by the World Health Organisation (WHO), defined as the symptoms that persist for more than three months after infection, and are of at least two months duration and not explained by an alternative diagnosis (Soriano et al., 2022). Conservative global estimates indicate that at least 10–20% of people who had COVID-19 develop LC (Nalbandian et al., 2023; National Center for Health Statistics, 2023), with numbers going up to 50–60% in cases that required hospitalisation (Chen et al., 2022; Pazukhina et al., 2022). In the United Kingdom alone, it is estimated that approximately two million people in private households are currently experiencing LC-symptoms (Office for National Statistics, 2023).

Long COVID is a complex condition characterised by a multitude of heterogenous symptoms including fatigue, shortness of breath, pain, cognitive disturbances, sleep problems and post-exertional malaise (Davis et al., 2021). The clinical manifestations of LC show similarities to the symptom phenotypes of other chronic conditions, namely Fibromyalgia Syndrome (FMS) and Chronic Fatigue Syndrome/Myalgic Encephalomyelitis (CFS/ME) (Bierle et al., 2021; Fialho et al., 2023). Fibromyalgia is a common chronic and widespread musculoskeletal pain condition, in which fatigue is also commonly reported. Individuals diagnosed with CFS/ME also present with pain, although the characteristic symptom of CFS/ME is persistent fatigue and post-exertion malaise. In both conditions, pain and fatigue are accompanied by many symptoms such as that sleep disturbance and brain fog that match those reported in LC. Unsurprisingly, recent studies show that a large proportion of LC patients fit the diagnostic criteria for FMS and CFS/ME (Bonilla et al., 2023; Haider et al., 2022; Ursini et al., 2021). At present, and much like LC, there is no diagnostic test or biomarker available for FMS or CFS/ME. Additionally, the three conditions have similar risk factors and demographics affecting more females than males with greater reporting in middle-aged individuals (Castro-Marrero et al., 2017; Goldhaber et al., 2022; Walitt et al., 2015).

The similar clinical picture of multisystem symptoms, and possible post-infectious nature of these conditions (Choutka et al., 2022), suggests they may have common underlying pathophysiological mechanisms. A mechanism known to play a major role in FMS and CFS/ME is central sensitisation (Bourke et al., 2015). This is characterised by maladaptive changes in excitability of central neurons that lead to amplification of central nervous system responses to a variety of inputs (Woolf, 2011). Central sensitisation provides an evidence-based explanation for symptoms that are often deemed medically unexplained (Latremoliere and Woolf, 2009; Smart et al., 2010). Quantitative electroencephalography (qEEG) is a reliable and valid non-invasive neuroimaging tool that has been extensively used to investigate functional alterations of the brain associated with central sensitisation in patients with FMS and CFS/ME (de Melo et al., 2021; Maksoud et al., 2020). In central sensitisation syndromes such as FMS and CFS/ME, there is an increased cortical responsiveness to afferent inputs involved in symptoms of pain, fatigue and other hypersensitivities commonly observed in patients.

Due to the novelty of the LC condition, the contribution of maladaptive plasticity of the central nervous system to development and maintenance of symptoms remains relatively unexplored in LC (Fernández-de-las-Peñas et al., 2022; Khoja et al., 2022). The lack of controlled data in LC has motivated the need to look at similar conditions. This systematic review aims to synthetise and appraise existing evidence of alterations in resting-state qEEG in individuals with LC, FMS and CFS/ME compared to healthy controls. Drawing on existing qEEG research on conditions similar to LC may help elucidate possible mechanisms of central sensitisation in LC and lay the groundwork for future qEEG studies in LC. Characterisation of the neurophysiological signatures of LC is of particular importance to inform on potential diagnostic markers and promising targets of non-invasive neuromodulation techniques.

## 2. Methods

### 2.1. Search strategy

The protocol for this systematic review was pre-registered on PROSPERO (CRD42022382079). The review was performed in agreement with the Preferred Reporting Items for Systematic Reviews and Meta Analyses (PRISMA) guidelines

(BMJ, 2021;372:n71). A literature search of relevant articles published between December 1994 and September 2023 was conducted within the following databases: MEDLINE (Ovid), Embase (Ovid), CINHAL, PsycINFO and Web of Science (via Clarivate Analytics). Each database was searched systematically using keywords and MeSH headings relevant to the conditions of interest (COVID-19; SARS-CoV-2; Coronavirus; Long COVID; Post-COVID; Chronic COVID; Post-acute COVID; long-haul COVID; fibromyalgia; myalgic encephalomyelitis; chronic fatigue syndrome; postviral syndrome) and study methodology (EEG; electroencephalography; electric encephalography; qEEG). The search strategy was adapted to each database as necessary. Only articles written in English and available in full-text were included. We also undertook manual searching on Google scholar and performed a through scanning of reference lists for relevant articles.

### 2.2. Study selection

All studies fulfilling the following criteria were included: (1) involve human participants aged 18 or older; (2) Fibromyalgia diagnosis confirmed using the American College of Rheumatology criteria (Wolfe et al., 2010); or CFS/ME diagnosis confirmed using Fukuda case definition (1994), Canadian Consensus Criteria (2003) or International Consensus Criteria (2011) (Carruthers et al., 2003; Carruthers et al., 2011; Fukuda et al., 1994); and LC diagnosis as outlined by NICE guidelines (NICE, 2022); (3) investigate changes in resting-state brain activity using qEEG methods compared to healthy controls. Case reports, reviews, editorials and preprints were excluded, as well as any purely interventional studies. Physiologic studies asking participants to perform tasks or involving stimuli of any type (e.g. event-related potential studies) were also excluded.

The Rayyan software tool was used to import all retrieved references and for screening against eligibility criteria. Following the removal of duplicates, two authors (BSP and AT) screened the titles and abstracts for any exclusion criteria. The pre-selected studies underwent full-text review to determine if they met inclusion criteria. Where there was disagreement, study eligibility was reassessed by a third reviewer (OK).

### 2.3. Data extraction

Study characteristics, EEG protocol, primary outcome measures and results were independently extracted from the selected articles by two reviewers (BSP and OK). Findings were reported for four typical EEG frequency bands: the low-frequency bands delta (1–4 Hz), theta (4–8 Hz) and alpha (8–13 Hz); and the high-frequency beta band (13–30 Hz). Some studies subdivide the alpha band into alpha-1 (8–10 Hz) and alpha-2 (10–13 Hz); and the beta band into beta-1 (13–18 Hz), beta-2 (18–22 Hz), beta-3 (22–30 Hz), although frequency ranges vary across studies.

### 2.4. Quality assessment

All articles selected for inclusion were critically appraised using the Newcastle-Ottawa Scale (Wells et al., 2000) to evaluate methodological quality and risk of bias. This instrument has separate assessment criteria for case-control and cohort studies. Critical appraisal was performed by two independent reviewers (BSP and OK), with any discrepancies resolved by consulting another co-author (AT). Each study was scored based on three domains using a star-rating system:

(1) Selection: Assigns a maximum of four stars for appropriate definition, representativeness and selection of study participants.
(2) Comparability: Assigns one star if the most important confounding factors were adjusted for in the analysis or study design, and an extra star for additional adjustment for any other confounding factors. For this review, age and sex were selected as the most important confounding factors.
(3) Exposure/outcome: Assigns a maximum of three stars for use of appropriate methods to ascertain exposure (for case-control studies) or evaluate outcome of interest (for cohort studies).

Total scores in the range of 0–3 indicate low quality, 4–6 moderate quality, and 7–9 high quality.

## 3. Results

A total of 2510 studies were identified through database searching on MEDLINE (838), CINAHL (205), EMBASE (398), PsycINFO (151) and Web of Science (918). Following duplicate removal and screening based on meeting the requirements established by the inclusion and exclusion criteria, the final number of articles included in the review was refined to 17 (PRISMA flow diagram in Figure 1). No additional relevant articles were identified through manual searching. Eight of the selected studies compared resting-state EEG collected from healthy controls with a sample of FMS patients (González-Roldán et al., 2016; González-Villar et al., 2020; Hargrove et al., 2010; López et al., 2015; Makowka et al., 2023; Martín-Brufau et al., 2021; Vanneste et al., 2017; Villafaina et al., 2019), six involved CFS/ME patients (Billiot et al., 1997; Flor-Henry et al., 2009; Sherlin et al., 2007; Wu et al., 2016; Zinn and Jason, 2021; Zinn et al., 2018) and the remaining three studies focused on LC (Cecchetti et al., 2022; Kopańska et al., 2022; Ortelli et al., 2023). Table 1 shows a summary of the methodology and findings of the selected studies.

**Figure 1.**
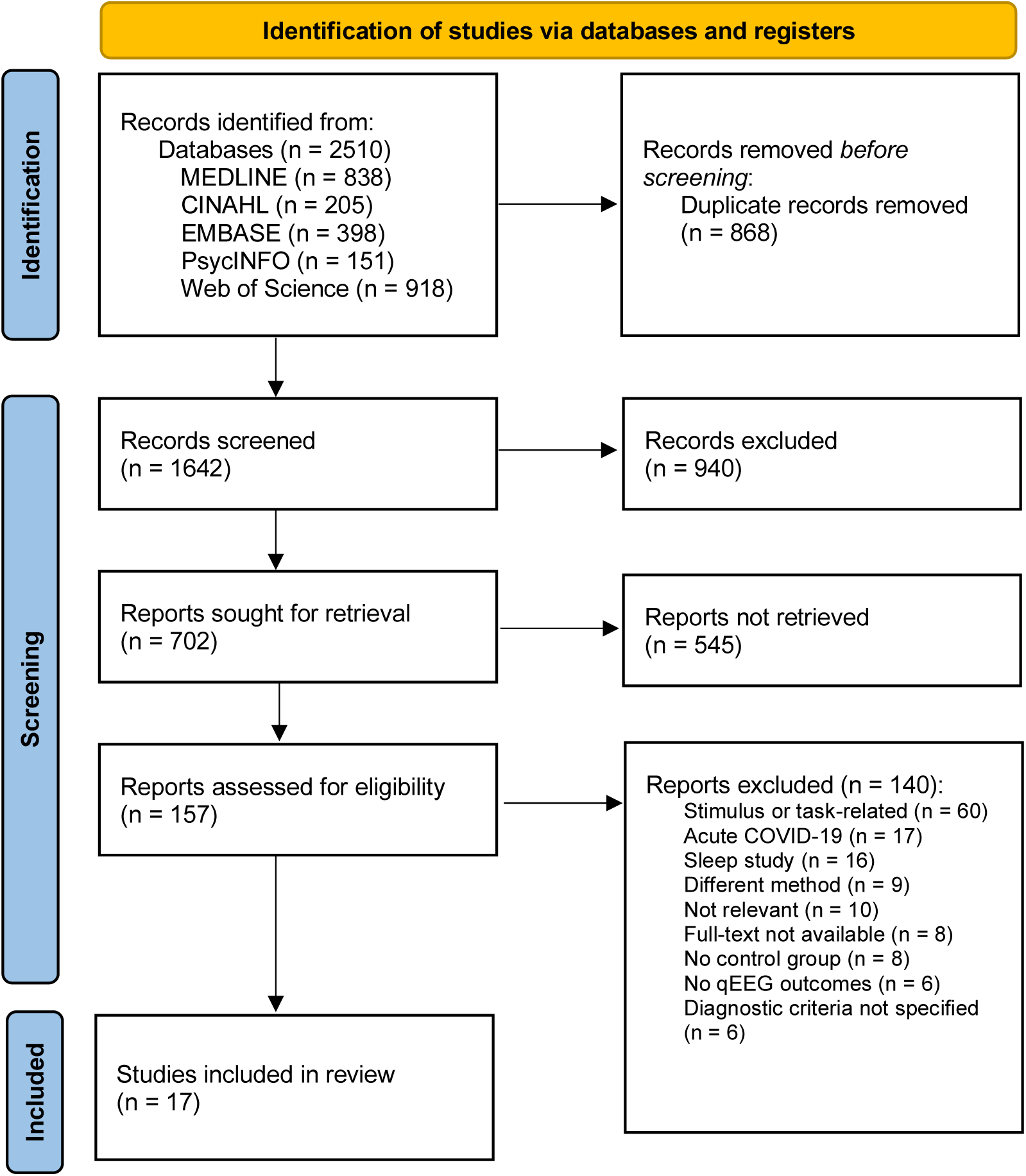
PRISMA flow diagram of screening and selection process.

**Table 1.**
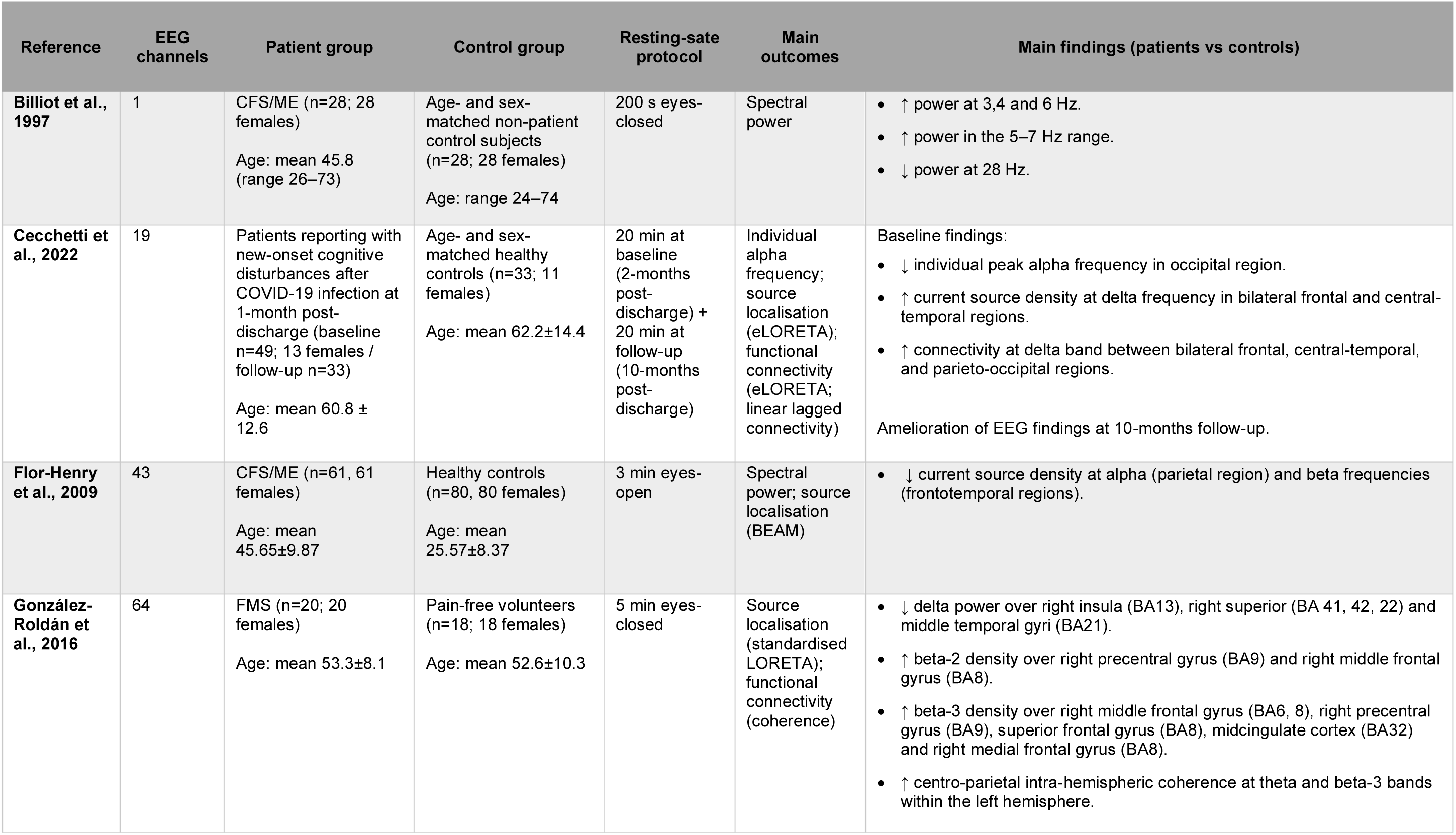
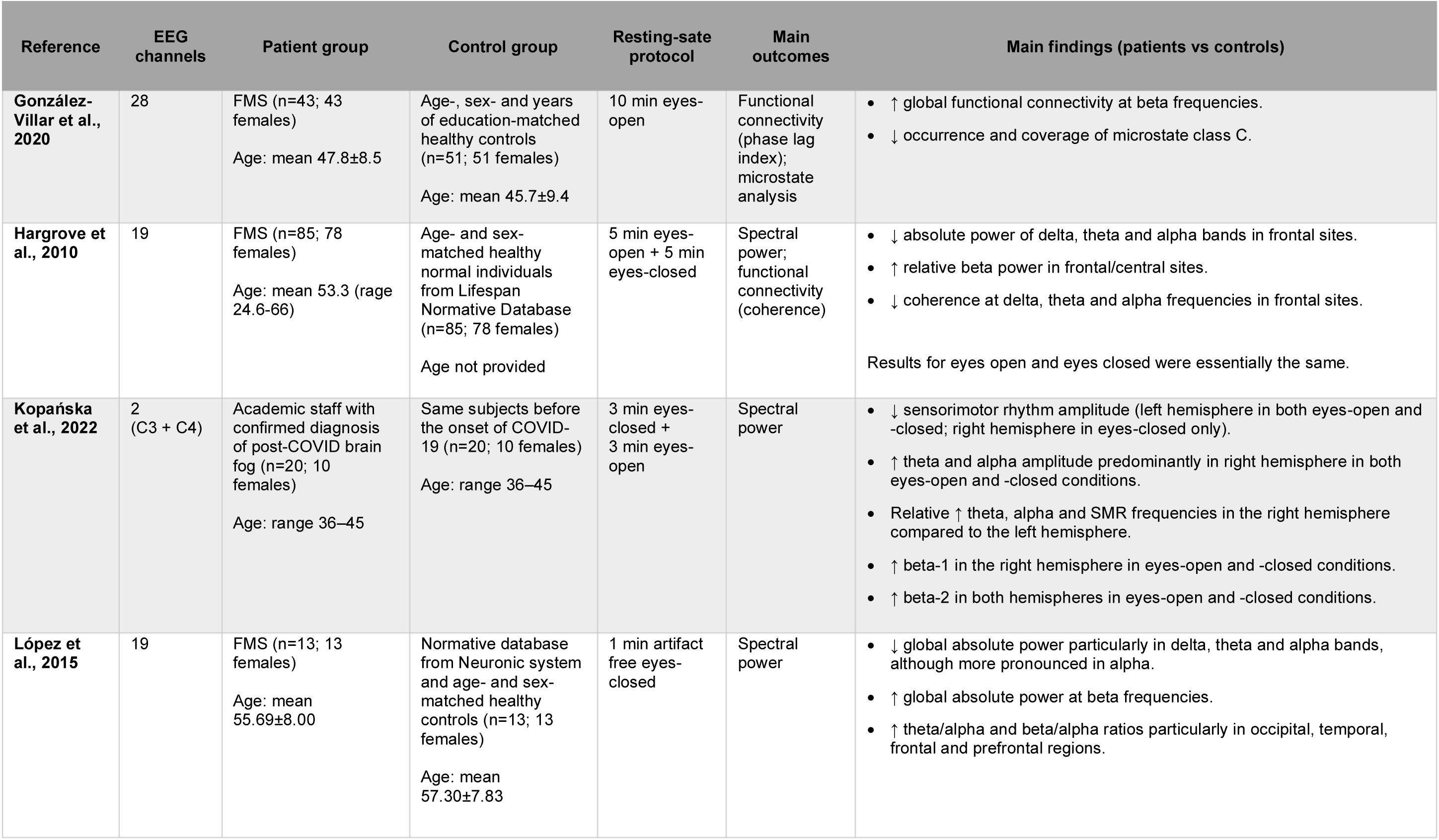
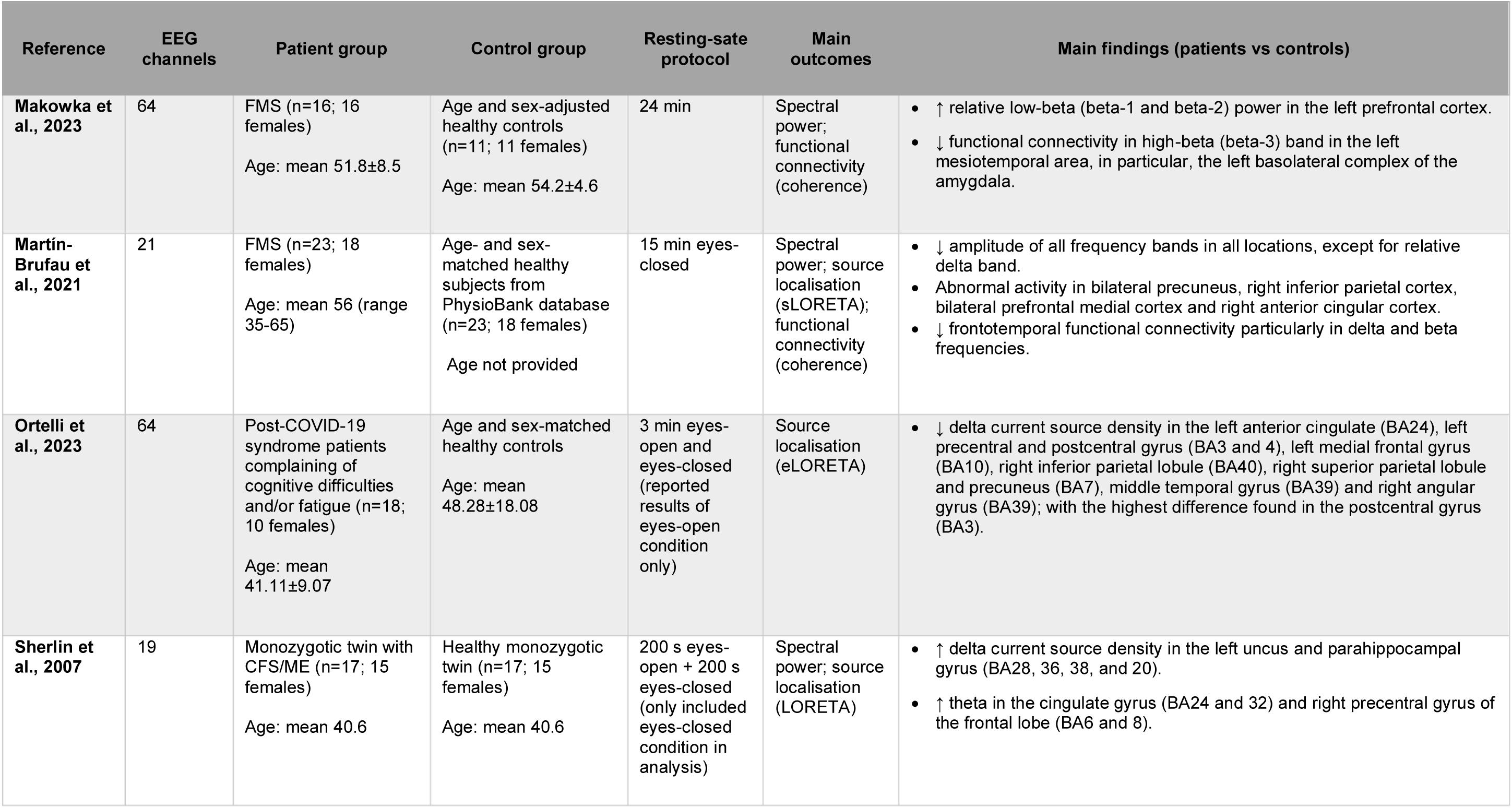
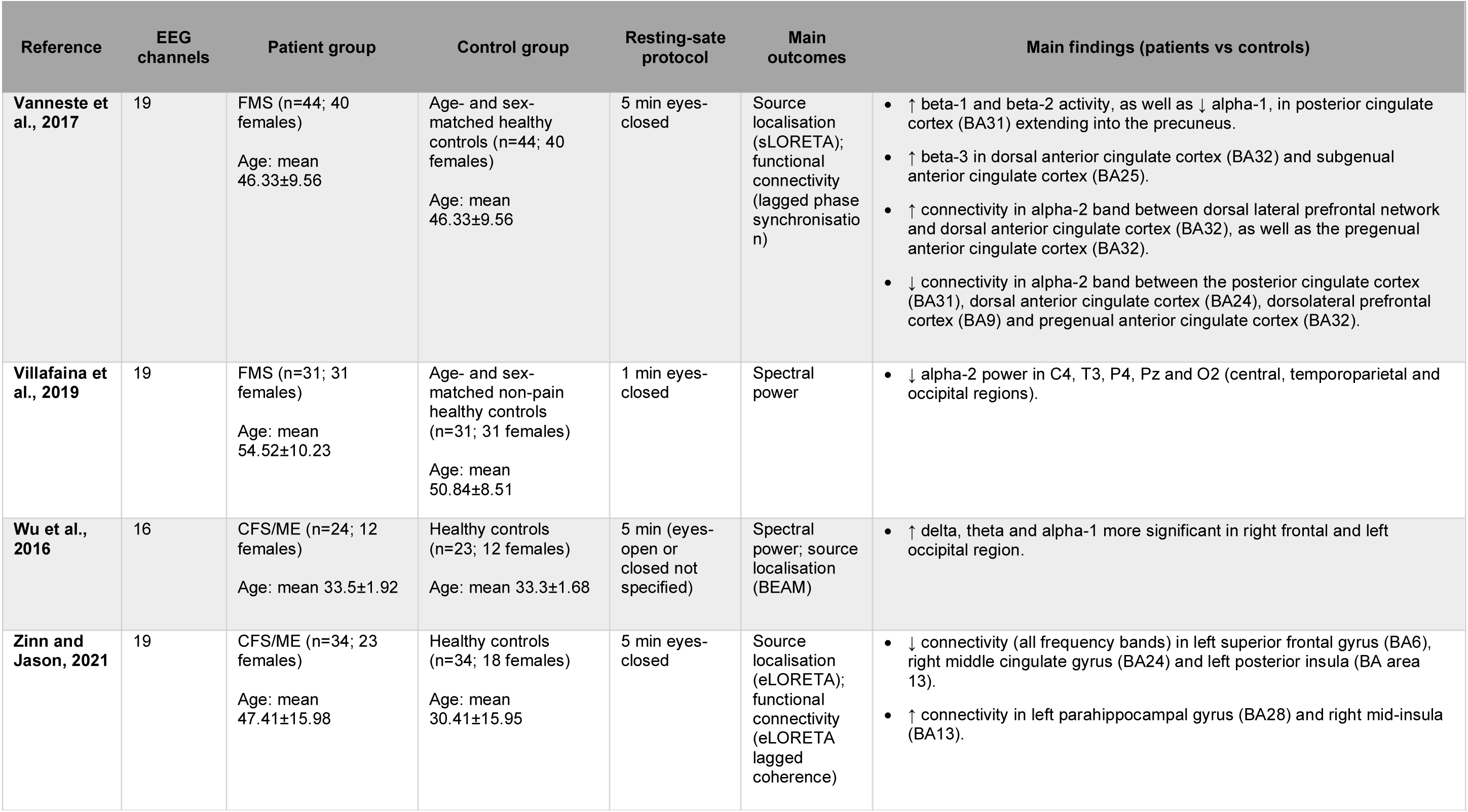
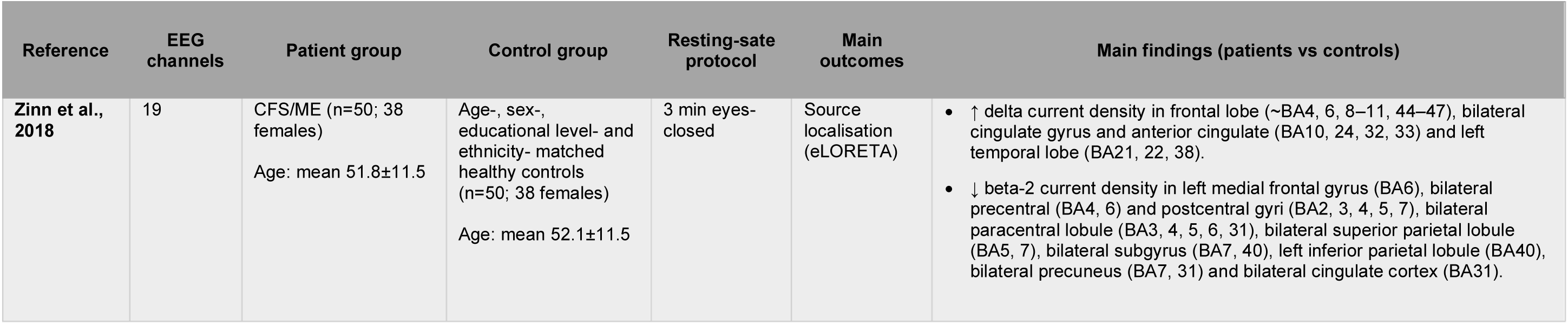
Description of EEG protocol, patient and control groups, and main findings from the selected studies.

### 3.1. Literature reporting changes in electrical activity of resting-state qEEG

We first report the findings from studies on the spectral characteristics (frequency and power) of resting-state EEG signals. Ten of the selected studies reported results of spectral analysis of resting-state EEG signals: five in FMS (Hargrove et al., 2010; López et al., 2015; Makowka et al., 2023; Martín-Brufau et al., 2021; Villafaina et al., 2019), three in CFS/ME (Billiot et al., 1997; Flor-Henry et al., 2009; Wu et al., 2016) and two in LC (Cecchetti et al., 2022; Kopańska et al., 2022). The findings described in this section that map changes in each frequency band to broad brain regions are represented in Figure 2 (scalp map in the centre of each panel).

**Figure 2.**
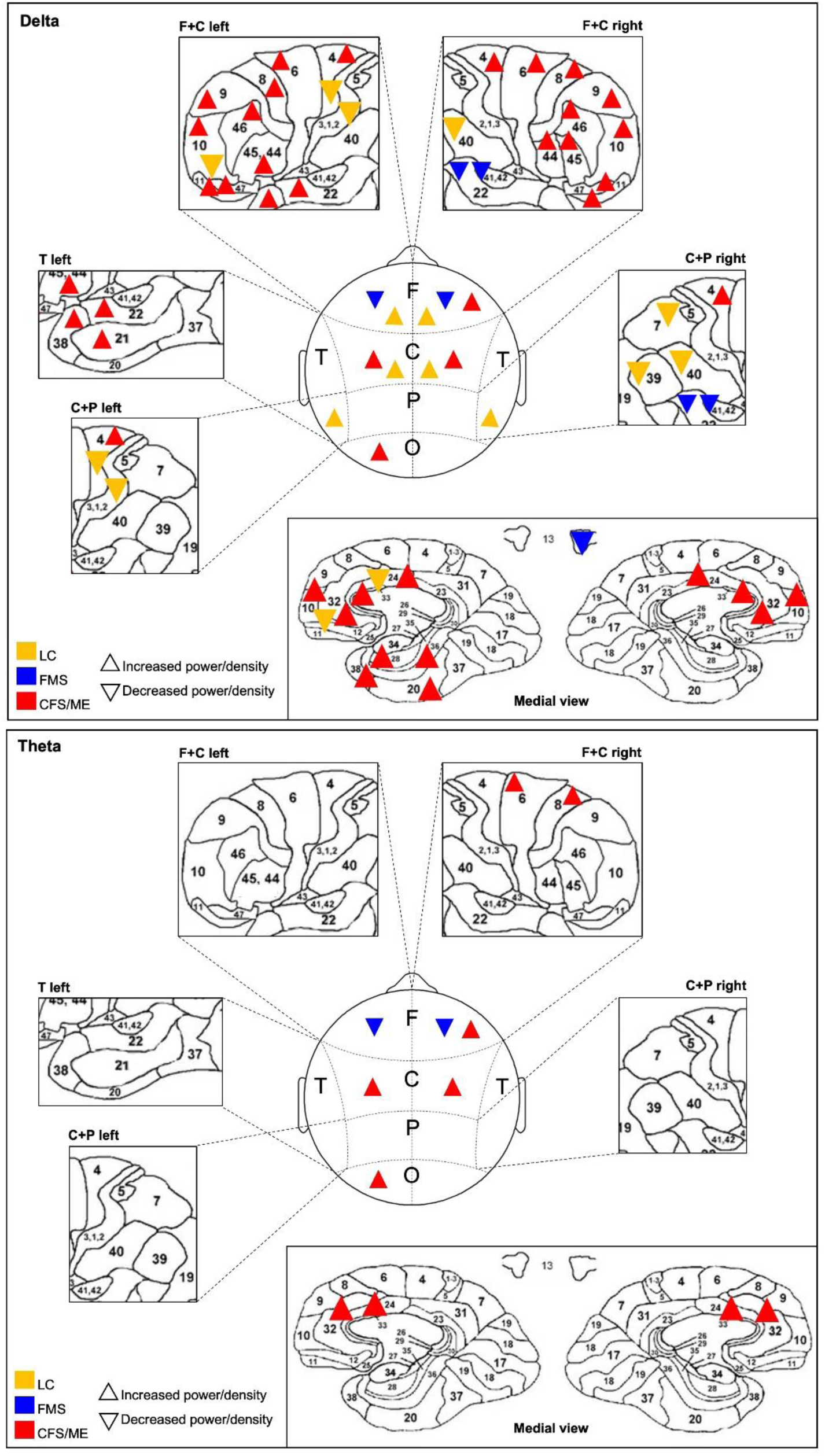
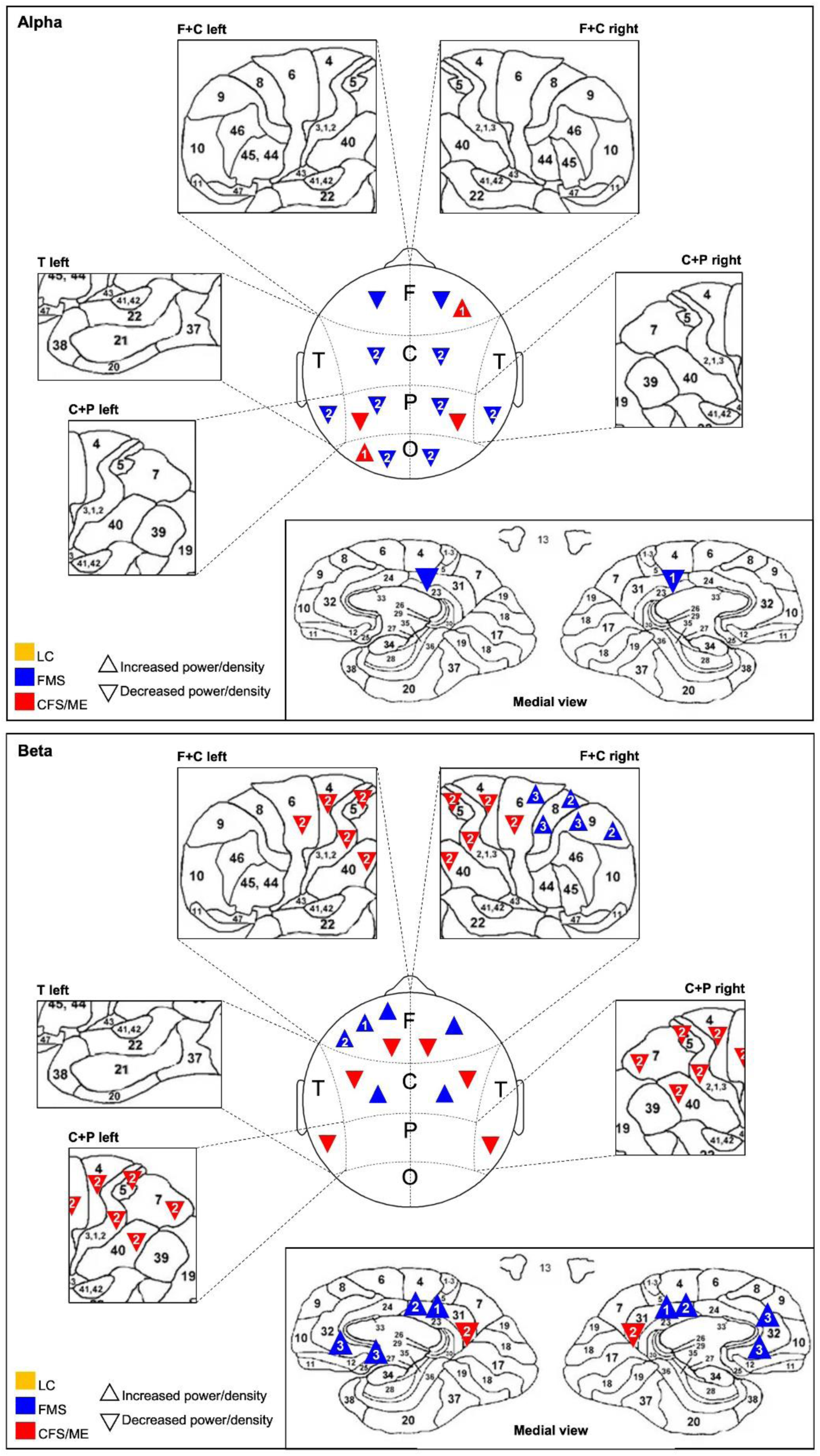
Spatial representation of the findings of the selected studies reporting changes in electrical activity of resting-state EEG signals in LC (yellow), FMS (blue) and CFS/ME (red) patients compared to healthy controls. Each panel represents the findings relative to each frequency band: delta, theta, alpha and beta. Scalp maps (in the centre of each panel) highlight the findings from studies reporting changes in broad brain regions: frontal (F), central (C), parietal (P) and temporal (T). The expanded windows are to highlight the findings from studies reporting changes in specific Brodmann areas. The window in the lower corner of each panel shows a medial view of the brain highlighting changes found in subcortical areas. Significant findings from each study are represented in the relevant brain region or Brodmann area by a triangle: pointing upwards () for increased power/current source density or pointing downwards () for decreased power/current source density compared to healthy controls.

All FMS studies found a significantly reduced activity of low-frequency bands compared to healthy controls (Figure 2). For instance, López et al. (2015) reported lower absolute power of delta, theta and alpha bands across all scalp regions; although more pronounced in alpha. Another study using EEG recordings of longer duration (15 min vs 1 min) showed similar findings for theta and alpha bands but not for delta, for which the difference between patient and control groups was not statistically significant (Martín-Brufau et al., 2021). In Hargrove et al. (2010), the significant decrease in delta, theta and alpha bands relative to matched controls from a normative database was limited to frontal sites. This was true for both eyes-open and eyes-closed resting-state conditions. Villafaina et al. (2019)’s study involving 31 females with FMS found significant differences only for alpha-2 power in electrodes located in central, temporoparietal and occipital regions. Regarding the higher-frequency EEG beta band, most studies in FMS reported a significant increase in beta power when compared to healthy controls (Hargrove et al., 2010; López et al., 2015; Makowka et al., 2023). The high relative beta power was observed in frontal and central EEG measurement sites in Hargrove et al. (2010), and generally across the scalp in López et al. (2015). Makowka et al. (2023)’s findings were limited to beta-1 and beta-2 sub-bands specifically in the left prefrontal cortex. One study found the opposite pattern, with FMS patients presenting with significantly decreased general beta activity compared to controls (Martín-Brufau et al., 2021).

In regard to CFS/ME, Wu et al. (2016) observed significantly increased values of delta, theta and alpha-1 power in CFS/ME patients compared to controls, particularly in right frontal and left occipital sites. Another study using single-channel EEG recorded significantly higher power at 3–7 Hz frequencies and significantly lower at 28 Hz, that fall within the range of delta/theta and beta bands, respectively (Billiot et al., 1997). Lastly, Flor-Henry et al. (2010) found a significantly decreased density at beta and alpha frequencies in CFS/ME patients compared to controls in the frontotemporal and parietal regions, respectively.

For LC, a study recording EEG signals from the same subjects before and after development and clinical confirmation of post-COVID brain fog found a relative increase in theta and alpha activity after COVID-19 infection (Kopańska et al., 2022). Moreover, the researchers noted a hemispheric lateralisation of the findings reflected by the relatively increased theta and alpha power in the right hemisphere compared to the left. Beta-1 power was found to be increased in the right hemisphere in both eyes open and closed conditions, while beta-2 power was increased in both hemispheres. No significant differences were found for the delta band. A longitudinal study found reduced baseline individual peak alpha frequency in the occipital region in patients with post-COVID cognitive disturbances compared to controls (Cecchetti et al., 2022). They also reported a significantly elevated baseline density at delta frequencies in frontal, central and temporal regions bilaterally in patients with post-COVID cognitive disturbances compared to controls.

### 3.2. Literature reporting changes in electrical activity mapped to Brodmann’s areas

In this section, we describe the results obtained from studies employing EEG source localisation methods that map changes in electrical activity of resting-state EEG to specific Brodmann areas (BAs). These studies offer a more granular understanding of spatial distribution of neural sources responsible for generating recorded EEG signals. Of the selected studies, five reported on specific BAs: two in FMS (González-Roldán et al., 2016; Vanneste et al., 2017), two in CFS/ME (Sherlin et al., 2007; Zinn et al., 2018) and one in LC (Ortelli et al., 2023). The findings described in this section are represented in Figure 2 (expanded windows of each panel).

Both studies on FMS reported increases in activity at beta band frequencies, particularly within the frontal cortex and cingulate regions (Figure 2). In Vanneste et al. (2017), significant differences were observed for beta-1 and beta-2 bands in the dorsal posterior cingulate (BA 31) and extending into the precuneus. The same was reported for alpha-1. Increased beta-3 activity was found in the dorsal and subgenual anterior cingulate cortex (BA32 and 25). A study using a similar protocol but a higher density EEG recording system also found increased density of beta-3 over the dorsal anterior cingulate cortex (BA32), along with significant increases in the right dorsolateral, medial and posterior regions of the frontal cortex (BA6, 8 and 9) (González-Roldán et al., 2016). Beta-2 density in the FMS group was significantly higher in BA8 and BA9 of the right hemisphere compared to the control group. Changes in delta activity were also observed, particularly a reduced power over the right insula and right superior and middle temporal gyri (BA13; 22, 41, 42; and 21, respectively).

The two studies on CFS/ME reported increased delta current density in multiple brain regions, which seem to contrast with the findings for FMS (Figure 2). In more detail, Sherlin et al. (2007) found significant differences in the left parahippocampal gyrus, anterior and inferior area of the temporal lobe (BA28, 36, 38 and 20, respectively) in CFS/ME patients compared to their healthy monozygotic twins. Similarly, Zinn et al. (2018)’s study including matched unrelated-healthy controls also showed an increase in delta in the left temporal lobe (BA21, 22, 38); as well as bilaterally in areas of frontal lobe and anterior cingulate cortex. They also noted a significant decrease in beta-2 in the dorsal posterior cingulate and precuneus (BA31) as opposed to Vanneste et al. (2017)’s observations in FMS; that extended to areas of the precentral gyrus (BA2, 3, 4), premotor cortex (BA6), superior parietal lobe (BA5, 7) and left inferior parietal lobe (BA40). Lastly, the abovementioned twin study highlighted a significant increase in current density within the theta band range in the anterior cingulate gyrus (BA24, 32) and right precentral gyrus (BA6, 8).

A LC study by Ortelli et al. (2023) found a decrease in delta current source density in the frontoparietal lobe bilaterally and the left temporal lobe in LC patients experiencing cognitive disturbances and/or fatigue compared to controls. In particular, they reported significant differences in the: left anterior cingulate (BA24), left precentral and postcentral gyrus (BA3 and 4), left medial frontal gyrus (BA10), right inferior parietal lobule (BA40), right superior parietal lobule and precuneus (BA7), middle temporal gyrus (BA39) and right angular gyrus (BA39); with the highest difference found in the postcentral gyrus (BA3).

### 3.3. Literature reporting changes in functional connectivity of resting-state qEEG

We outline the findings derived from functional connectivity analysis of resting-state EEG signals in this section, providing insights into network dynamics and functional interactions between brain regions. EEG functional connectivity was assessed in eight studies: six in FMS (González-Roldán et al.,2016; González-Villar et al., 2020; Hargrove et al., 2010; Makowka et al., 2023; Martín-Brufau et al., 2021; Vanneste et al., 2017), one in CFS/ME (Zinn and Jason, 2021) and one in LC (Cecchetti et al., 2022).

The studies in FMS yielded inconsistent findings on differences between brain functional connectivity in patients and healthy controls. González-Roldán et al. (2016) found significant differences within the left hemisphere only, specifically an increased centro-parietal coherence at theta and beta-3 bands in the FMS group compared to pain-free controls. This contrasts with the observations by Makowka et al. (2023) of reduced functional connectivity at beta-3 band in areas of the left mesiotemporal area, in particular the basolateral complex of the amygdala. Another study reported reduced functional connectivity at low-frequency EEG bands (delta, theta and alpha) in frontal sites compared to normative database (Hargrove et al., 2010). Similarly, Martín-Brufau et al. (2021) found reduced functional connectivity in delta frequencies as well as in high-frequency beta band, particularly between frontal and temporal brain regions. Vanneste et al. (2017) identified changes related to alpha-2 band connectivity in FMS patients relative to matched-controls. Connectivity in the alpha-2 band was found to be reduced between the right dorsal lateral prefrontal cortex and bilateral insula, as well as between the right dorsal lateral prefrontal cortex and bilateral pregenual and dorsal anterior cingulate cortex. The opposite was found for connections within different regions of the cingulate cortex and regarding connectivity between the right insula and the left insula, bilateral posterior, dorsal anterior and pregenual cingulate cortex, where alpha-2 connectivity was increased. No significant differences were found for other frequency bands. A study by González-Villar et al. (2020) showed global effects on EEG connectivity, with the FMS group showing higher beta band connectivity on average compared to the control group.

For CFS/ME, Zinn and Jason (2021) showed reduced connectivity in the left superior frontal gyrus (BA6), right ventral anterior cingulate cortex (BA24) and left posterior insula (BA13) across all frequency bands in patients. They also observed increased connectivity in the left parahippocampal gyrus (BA28) and right insula (BA13).

Adding to the qEEG outcome measures outlined previously, the longitudinal study on post-COVID cognitive disturbances by Cecchetti et al. (2022) also analysed brain functional connectivity. Baseline resting-state EEG signals showed higher connectivity at delta frequencies in bilateral frontal, central-temporal, and parieto-occipital regions in the patient group compared to controls.

### 3.4. Literature reporting microstate analysis of resting-state qEEG

Lastly, we present the findings of studies reporting on EEG microstates. EEG microstate analysis offers a unique perspective on the temporal dynamics and discrete functional states of brain activity.

Only one study reported the outcomes of microstate analysis of resting-state EEG signals. González-Villar et al. (2020) found a decrease in the occurrence and coverage of microstate class C in a sample of FMS patients when compared to healthy matched controls. No microstate studies were identified in CFS/ME or LC conditions.

### 3.5. Quality assessment of the selected studies

The mean score of quality assessment of the selected studies was 6.47, indicating a moderate to high quality. The Newcastle-Ottawa Scale score for each study is shown in Table 2. Regarding the selection domain, recruitment and selection of the healthy control group was insufficiently described in the majority of the studies. Cases and controls were matched for sex and/or age in all studies, except in Wu et al. (2016) and Zinn and Jason (2021).

**Table 2.** Newcastle-Ottawa Scale quality assessment scores for each selected study.

| Reference | Study design | Newcastle-Ottawa Scale's Quality criteria |  |  | Total score |
| --- | --- | --- | --- | --- | --- |
|  |  | Selection | Comparability between groups | Exposure/ Outcome |  |
| Billiot et al., 1997 | Observational case-control | *** | * | ** | 6* |
| Cecchetti et al., 2022 | Longitudinal cohort | ** | * | *** | 6* |
| Flor-Henry et al., 2009 | Observational case-control | *** | * | ** | 6* |
| González-Roldán et al., 2016 | Observational case-control | **** | * | ** | 7* |
| González-Villar et al., 2020 | Observational case-control | *** | ** | ** | 7* |
| Hargrove et al., 2010 | Observational case-control | *** | * | ** | 6* |
| Kopańska et al., 2022 | Longitudinal cohort | *** | ** | *** | 8* |
| López et al., 2015 | Observational case-control | *** | * | ** | 6* |
| Makowka et al., 2023 | Observational case-control | **** | * | ** | 7* |
| Martín-Brufau et al., 2021 | Observational case-control | *** | * | ** | 6* |
| Ortelli et al., 2023 | Observational case-control | **** | * | ** | 7* |
| Sherlin et al., 2007 | Observational twin study | **** | ** | ** | 8* |
| Vanneste et al., 2017 | Observational case-control | *** | * | ** | 6* |
| Villafaina et al., 2019 | Observational case-control | *** | * | ** | 6* |
| Wu et al., 2016 | Observational case-control | *** | -- | ** | 5* |
| Zinn and Jason, 2021 | Observational case-control | **** | -- | ** | 6* |
| Zinn et al., 2018 | Observational case-control | *** | ** | ** | 7* |

## 4. Discussion

The primary aim of this systematic review was to synthesise and appraise the literature on resting-state qEEG patterns in three similar clinical syndromes: LC, FMS and CFS/ME. Seventeen studies were identified, all reporting significant differences in resting-state EEG signals in patients compared to healthy controls. Evidence of qEEG abnormalities in LC is scarce, limited to three studies focusing mainly on cognitive symptoms. The included studies on LC show inconsistent findings. The synthesis of the reviewed studies on FMS generated two frequent observations: (1) markedly decreased activity of low-frequency bands in patients, particularly within the alpha frequency range; and (2) increased activity of high-frequency band beta. In contrast to observations in FMS, the patterns that emerged in CFS/ME appear to follow an opposite trend with a tendency for marked increases in activity of low-frequency bands, specifically delta and theta bands, coupled with lower activity levels in the beta frequency range. Findings of abnormal activity of the four main EEG frequency bands were reported in different brain regions.

### 4.1. Comparative findings in FMS and CFS/ME

Despite the significant overlap in symptomology, FMS is dominated by widespread pain while for CFS/ME the core symptom is severe fatigue. The distinct abnormal resting-state qEEG signatures observed in FMS and CFS/ME may relate to core differences in clinical features.

Alpha oscillations are known to reflect deactivation of brain regions and to be more pronounced in states of relaxation where the individual is alert but not engaged in any external stimulus/task and attention is turned internally (Klimesch et al., 2007; Nunez et al., 2001). It is becoming increasingly evident that alterations in the alpha frequency band are linked to pain chronicity, with a number of reports showing an association between reduced alpha power and pain-related outcomes in chronic pain patients (Camfferman et al., 2017; Zebhauser et al., 2023). Our findings of abnormally reduced alpha power in FMS support the negative association between pain and EEG alpha rhythms observed in existing chronic pain literature. The direction of changes in resting-state EEG alpha activity in CFS/ME is not as well established, with the only two studies reporting alterations in alpha power in CFS/ME presenting conflicting findings (Flor-Henry et al., 2010; Wu et al., 2016).

Although established in the sleep literature, the functional significance of slow-wave delta, typically predominant during deep restorative sleep, is not well-defined in awake EEG. In CFS/ME, the elevated delta power reported in four of the selected studies could indicate the persistence of sleep EEG features during wakefulness, as proposed by previous studies on insomnia patients (Wu et al., 2013; Zhao et al., 2021). This in conjunction with an increase in theta power may reflect a general state of cortical hypoarousal or EEG slowing that supports the overwhelming feelings of mental fatigue and difficulty in maintaining a fully alert state experienced by CFS/ME patients.

This review found a tendency for decreased theta power in FMS with two studies reporting global effects and one reporting a regional decrease in frontal theta power (Hargrove et al., 2010; López et al., 2015; Martín-Brufau et al., 2021). Interestingly, this finding contrasts with the literature on other chronic pain conditions that reports increases in theta power compared to healthy controls (Sarnthein et al., 2006; Stern et al., 2006). This has been previously noted by Pinheiro et al. (2016) who argued that the different EEG signatures may reflect condition-specific pathophysiology in FMS compared to chronic neuropathic pain conditions.

As for beta oscillations, activity within this frequency range is associated with cognitive processing typically increased when the individual is active and concentrated, but especially in situations known to induce stress and anxiety (Engel and Fries, 2010; Spitzer and Haegens, 2017). The tendency to abnormally increased high-frequency beta activity found in the selected studies on FMS may be indicative of a state of central hyperexcitability that contrasts with that found in CFS/ME. Cortical hyperarousal in FMS may be associated with ongoing cognitive processing, possibly linked to rumination, along with increased levels of emotional distress and alertness characteristic of FMS.

While an overall tendency for increased or decreased levels of activity in each frequency band could be identified for FMS and CFS/ME, one should note that these observations were reported in distinct brain regions across the selected studies using source localisation and functional connectivity qEEG analysis methods. It is beyond the scope of this review to delve into the detailed functional implications of altered qEEG activity within each brain region and between brain regions. Notwithstanding, it is of particular relevance to highlight the overlap in brain regions implicated in the findings of disrupted oscillatory activity and functional connectivity in FMS and CFS/ME. Brain regions consistently reported include areas of the prefrontal cortex, anterior and posterior cingulate cortex and insula that are central hubs of two main brain networks: the default mode network and the salience network (Greicius et al., 2003; Seeley et al., 2007). The default mode network, active mainly at rest during self-referential processes such as mind-wandering and inward reflection, and the salience network interact dynamically to regulate the shift in allocation of attention and information processing to salient internal or external stimuli (Goulden et al., 2014). Disruptions in oscillatory activity and/or functional connectivity between regions of these networks could reflect an imbalance between the default mode network and the salience network in both conditions, which may be functionally relevant to impairments in cognitive, sensory and emotional processing in patients. Moreover, the overlap in brain regions could be suggestive of an interplay of central mechanisms that modulate the interconnected experience of pain and fatigue in FMS and CFS/ME.

### 4.2. Preliminary findings in LC

To date, only three studies assessed resting-state qEEG in individuals with LC compared to healthy controls, all of which identified spectral power changes in patients. Notwithstanding, findings were inconsistent across the selected studies. This is particularly true for the delta frequency band, with reports of excessively high (Cecchetti et al., 2022) or excessively low delta activity (Ortelli et al., 2023), as well as no significant differences (Kopańska et al., 2022) with respect to controls. The mixed results could reflect LC cohort being a heterogeneous group with different patients having different dominant symptom patterns (phenotypes). For instance, Cecchetti et al. (2022) included only individuals that required hospitalisation during the acute phase of COVID-19 infection and self reporting post-COVID cognitive impairments. Their findings of heightened delta activity in LC patients agree with common reports of EEG slowing in severely ill COVID-19 patients (Kubota et al., 2021). Ortelli et al. (2023), on the contrary, included only non-hospitalised LC patients complaining of cognitive impairments and/or fatigue. The contrasting qEEG patterns could be attributable to the varying degrees of severity of acute COVID-19 infection, in addition to possible intensive care-induced effects on the central nervous system.

Although no general trends could be inferred for LC due to limited study availability and heterogeneity, the early findings of qEEG abnormalities described in this review support a possible role for central sensitisation in LC patients. This adds to a growing body of neuroimaging research suggesting both structural and functional alterations of the central nervous system in LC patients (Chang et al., 2023; Díez-Cirarda et al., 2023; Voruz et al., 2022). Nevertheless, the qEEG patterns observed in the selected LC studies do not seem to follow the trends described in FMS or CFS/ME. It should also be noted that currently available resting-state qEEG studies focus mainly on cognitive symptoms of LC, not drawing on other neurological symptoms of LC such as pain and fatigue, and thus cannot be generalised to the LC population. Taking previous research on similar chronic conditions into consideration, it would be interesting to investigate whether LC patients presenting with phenotype more closely resembling FMS (chronic pain being the leading symptom) show qEEG patterns more similar to those associated with FMS. The same could also be applied to LC patients presenting mostly with fatigue-dominant phenotype and resemblance to qEEG abnormalities observed in CFS/ME.

### 4.3. Limitations

This systematic review has several limitations. While all included papers were rated moderate to high quality on the Newcastle-Ottawa Scale, the studies were limited to small sample sizes. Methodology varied widely across studies, with each study employing different sample matching processes, resting-state protocols (eyes open versus closed), recording systems (single-channel to high-density 64-channel EEG), and EEG processing and analysis techniques. Moreover, in studies employing methods of EEG source localisation and functional connectivity, the process of selection of regions of interest may have introduced bias in the analysis, hence affecting comparability of the results. It is also important to consider the borderline between FMS and CFS/ME, and now also LC, is a subtle one as these conditions commonly co-occur. This could further contribute to variability and inconsistencies in results. We were unable to perform meta-analysis due to heterogeneity in study methods and reported qEEG outcome measures. This systematic review is also limited in that it did not extract and interpret data regarding correlations between qEEG features and severity of symptoms, or other variables collected by the selected studies.

### 4.4. Recommendations for future research

This is the first systematic review of resting-state qEEG studies in LC and similar chronic conditions. Expanding upon the findings of previous research on FMS and CFS/ME, we suggest that phenotyping LC patients based on predominant symptoms, in particular pain-dominant and fatigue-dominant subgroups, may reveal EEG patterns analogous to those associated with FMS and CFS/ME, respectively. Future qEEG research should focus on capturing the multifaceted nature of LC by measuring the severity of all common neurological symptoms associated with the condition and acknowledging the diverse LC phenotypes. Emphasis should also be placed on standardising EEG recording protocols and analysis methods, whenever possible, to improve comparability of findings. Microstate analysis should be considered in future studies as alterations in microstate dynamics remains largely unexplored in these conditions. Characterisation of the EEG signatures of LC will help uncover the mechanisms of central sensitisation underlying medically unexplained neurological symptoms of LC. Emerging evidence could support the use of qEEG as a reliable diagnosis and monitoring tool for LC, and translate into new therapeutic targets for non-invasive neuromodulation therapies tailored to the different LC phenotypes. Because of the many similarities between LC, FMS and CFS/ME, advances in LC research could also be relevant to other poorly understood central sensitisation syndromes.

## 5. Conclusions

Our findings suggest that the qEEG patterns associated with FMS and CFS/ME follow opposite trends in terms of activity of the different brainwave bands. Further research is required to provide more conclusive evidence on the similarities and differences between qEEG signatures of different LC phenotypes and central sensitisation syndromes similar to LC.

## Data availability statement

Data will be made available on request.

## Author contributions

BSP, MS, CB, ID and AJC contributed to the conception and design of the study. BSP and AT conducted the systematic literature search and screening. BSP and OK performed data extraction and critical appraisal of the selected studies. MS provided clinical input as a clinician managing patients with the conditions in his practice. All authors contributed to interpretation of the data. BSP drafted the manuscript. All authors critically revised the manuscript and approved the version for publication.

